# Relationships Between Parental Involvement and Suicidal Ideation among In-school Adolescents in Vietnam: A Multilevel Analysis of the Global School-Based Student Health Survey 2019

**DOI:** 10.1101/2021.03.11.21253432

**Authors:** Khuong Quynh Long, Tran Thi Tuyet Hanh, Hoang Thi Ngoc Anh, Kidong Park, Momoe Takeuchi, Nguyen Tuan Lam, Pham Thi Quynh Nga, Le Phuong Anh, Le Van Tuan, Tran Quoc Bao, Nguyen Hang Nguyet Van, Pham Quoc Thanh, Nguyen Hong Phi, Le Duong Minh Anh, Nguyen Hong Phuong, Hoang Van Minh

**Affiliations:** Hanoi University of Public Health, Hanoi, 100000, Vietnam; World Health Organization, Country Office for Viet Nam, Hanoi, 100000, Viet Nam; Vietnam Ministry of Education and Training, Hanoi, 100000, Vietnam; General Department of Preventive Medicine, Ministry of Health, Hanoi, 100000, Vietnam; High school for Gifted student, Hanoi University of Science, Vietnam National University, Hanoi, 100000, Vietnam; Poverty, Health and Nutrition Division, International Food Policy Research Institute (IFPRI), Washington, DC 20006

**Keywords:** Suicidal ideation, Parental involvement, Adolescent, GSHS, Vietnam, Multilevel analysis

## Abstract

Youth suicide is a leading cause of death among adolescents, but evidence about the influences of parental involvement on adolescent suicidal behaviors is inconsistent and have not been well studied. We used nationally representative data from the Vietnam Global School-based Student Health Survey (GSHS) 2019 (n = 7796 students aged 13–18 years). Using the 2-level random intercept logistic regressions, we evaluated the relationship between parental involvement (high expectation, monitoring, and understanding) and suicidal ideation and identified related factors of suicidal ideation. The overall prevalence of suicidal ideation was 15.6%. While high level of parental monitoring and understanding were associated with lower odds of suicidal ideation among adolescents (OR: 0.63; 95% CI: 0.52–0.77 and OR: 0.60; 95% CI: 0.49–0.73, respectively), high parental expectation was linked to higher odds of suicidal ideation (OR: 1.42; 95% CI: 1.24–1.63). Other risk factors at the individual- and school-level for suicidal ideation included being girls, living in urban areas, having mental health problems, involving in risk behaviors, suffering from bullying and violence, and poor school quality. Targeted suicide prevention initiatives should take into account comprehensive aspects of parent-child bonding, student, and school factors to mitigate the burden of suicidal behaviors among adolescents.

## Introduction

Youth suicide is a major public health concern worldwide and a leading cause of death among adolescents (Patton et al. 2009). It is estimated that about 164,000 self-inflicted deaths worldwide each year in individuals younger than 25 years old (Patton et al. 2009). Furthermore, about 75% of global deaths by suicide occur in low- and middle-income countries (LMICs), where 90% of children and young adults in the world reside (World Health Organization). A recent systematic analysis of 59 LMICs estimated that the prevalence of suicidal ideation is 16.9%, suicide plan 17.0%, and suicide attempts 17.0% in the last 12 months among adolescents aged 13-17 years (Uddin et al. 2019). The current rate might be under-recorded since youth suicide is often a sensitive topic (Gosney and Hawton 2007). Since adolescence is a vulnerable period for suicidal behaviors (Bilsen 2018), identifying risk factors for suicidal behaviors plays a crucial role in formulating public health strategies and policies.

The reasons behind suicidal behaviors are complex and affected by a variety of risk factors at multiple levels. At the individual level, these factors include demographic characteristics: higher age, a few close friends (Peltzer and Pengpid 2017; Putra et al. 2019; Uddin et al. 2019); mental health disorders: depression, anxiety, poor self-esteem, loneliness (Itani et al. 2017; Putra et al. 2019; Shayo and Lawala 2019; Wilson et al. 2012); lifestyle risk behaviors: drugs use, drinking, smoking (Almansour and Siziya 2017; Itani et al. 2017; Peltzer and Pengpid 2015; Putra et al. 2019); peer victimization: being violence, bullying victim, involved in the physical fight (Almansour and Siziya 2017; Dema et al. 2019; Page and West 2011); and poor academic performance (Almansour and Siziya 2017). At the household level, parental separation, parental divorce, or poor family relationships are the key factors associated with suicidal behaviors (Hawton et al. 2012; McMahon et al. 2010). In contrast, the availability of peer kindness and social support in surrounding enviroments protects adolescents against suicidal behaviors (Kerr et al. 2006; McMahon et al. 2010).

While evidence has been established on the association between the above-mentioned factors and youth suicide, the roles of parent-child bonding have not been well studied and are inconsistent. Previous studies found that adolescents connected to their parents were less likely to commit suicide (Kidd et al. 2006), and parental support and supervision were linked to low suicidal ideation and attempt (Dema et al. 2019; Peltzer and Pengpid 2017; Putra et al. 2019). However, the recent study using data from 48 LMICs indicated that parental over-protection (i.e., too much parental care and monitoring) was associated with higher odds of suicidal ideation and suicide plan among adolescents (Kim 2019).

Vietnam is a developing country in South-East Asia, with a population of 97 million people and more than 8 million in-school adolescents (Vietnam General Statistics Officice 2020). The Vietnam Global School-based Student Health Survey (GSHS) in 2013 reported that the prevalence of suicidal ideation was 16.9% (13.6% among senior secondary school and 18.6% among high school students) (Nguyen et al. 2019). However, there is a lack of up-to-date national representative data about suicidal behaviors and its related factors. The lack of such information may result in limited evidence to formulate strategies and policies to improve adolescent health in Vietnam. Moreover, Vietnam is a country with a traditional family-oriented culture where individuals are embedded components of a system rather than independent agents, and the multigenerational interaction (especially in rural areas) is more prominent in individuals’ lives. To our knowledge, no study has examined the relationship between parental involvement and adolescent suicidal behaviors in Vietnam.

To fill the literature gaps, we use the most recent data from Vietnam GSHS 2019 to: 1) examine the relationships between parental involvement and suicidal ideation among in-school adolescents, and 2) identify multilevel factors of suicidal ideation among adolescents using the multilevel analysis approach.

## Methods

### Study setting and participants

We used data from the Vietnam GSHS 2019. This survey used the standardized protocol of the WHO and US CDC, which was designed to collect data from school-aged adolescents in several developing countries (US Centers for Disease Control and Prevention). GSHS 2019 used a cross-sectional two-stage cluster sampling design to select a representative sample of students aged 13- 17+ that is equivalent to grades 8, 9 (senior secondary school), and 10, 11, 12 (high schools) in Vietnam. The GSHS covers key areas of behavioral risk factors and protective factors among adolescents (US Centers for Disease Control and Prevention), using a set of core questionnaire modules that address the leading causes of poor health and mortality, including suicidal thoughts and behaviors.

### Questionnaire development

We translated the current version of the English questionnaire (US Centers for Disease Control and Prevention) using the standardized forward and backward translation. We also used relevant indicators from the 25 Global Non-Communicable Diseases (NCDs) and Healthy Vietnam (World Health Organization) to adapt to the Vietnam GSHS 2019 questionnaire. We then piloted the translated questionnaire with 120 students aged 13–17 years from one secondary school and one high school to test for the comprehension of questions, the appropriateness of language, and any logical issues. The pilot data were reviewed by senior researchers to further revise the questionnaire for improving clarity. Students from these two schools were excluded from the final sample.

### Sample size and sampling procedures

The sample size was calculated to estimate the prevalence of each component of GSHS. In order to represent subpopulations, we stratified the sample by residential areas - rural/urban and school levels - senior secondary/high school. The minimum sample needed to get the marginal error of 5% for each stratum was 1500. Based on the substantial response rate of the Vietnam GSHS 2013 (96%) (Kim 2019), we chose the response rate of 90% for 2019 Vietnam GSHS. Therefore, the number of students needed for each stratum was 1667, and a total of 6668 respondents for four strata.

The 2019 Vietnam GSHS was conducted in 20 provinces and cities in Vietnam (Supplemental figure 1). We recruited participants using the two-stage cluster sampling procedure. In the first stage, we selected schools based on a probability proportional to enrollment size. In the second stage, for each selected school, we chose classes using the simple random sampling technique - two classes for each secondary school (grades 8 and 9) and three classes for each high school (grades 10, 11, and 12). All students in selected classes were eligible to participate.

In each selected class, a homeroom teacher supported us to explain to students about the study. We provided information about GSHS and a consent form for students and requested them to share these documents with their parents for signing as an agreement to participate in the study. The following day, we administered GSHS questionnaire among students who had consent from parents and were willing to participate in the study. Participants completed a self-administered questionnaire and marked their responses on a separate computer scan-able answer sheet. In total, we collected data from 7796 students in 81 schools and 210 classes. The school response rate of 96.4% and the student response rate of 97.0% made up the overall response rate of 93.5%.

### Conceptual framework

We were not able to find any comprehensive frameworks to explain potential associations between parental involvement and adolescent suicidal ideation in Vietnam. Therefore, we developed an evidence-based conceptual framework based on literature review to guide our analysis (Figure 1). We searched on Pubmed to find publications on factors related to adolescent suicidal behaviors. We used the keywords (“suicide” OR “suicidal” OR “self-harm”) AND (“adolescent” OR “teenager” OR “youth”) AND (“factor” OR “determinant” OR “association”) to identify articles published in English between 2010 and 2020. From these studies, we identified multilevel factors that could affect adolescent suicidal ideation, including the parental involvement, six groups of student-level factors (demographic characteristics, peer relationship, mental health, peer victimization, lifestyle risk behaviors, other factors), and the school-level factors.

**Fig. 1.**
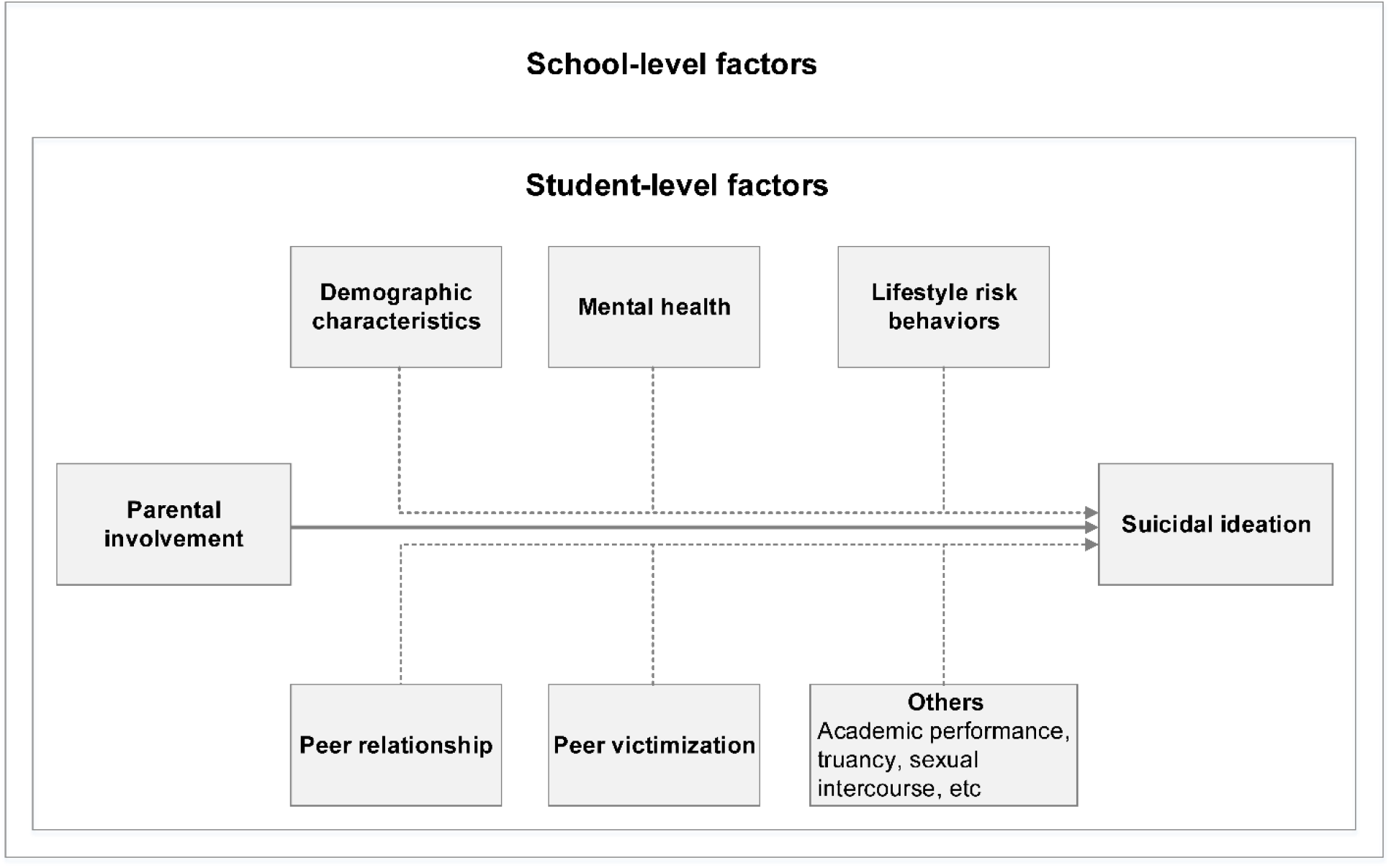
Conceptual framework of the relationship between parental involvement and suicidal ideation among in-school adolescents in Vietnam.

### Variables

We constructed variables based on the above-mentioned conceptual framework. The variable definitions in this study are provided in Table 1.

**Table 1.**
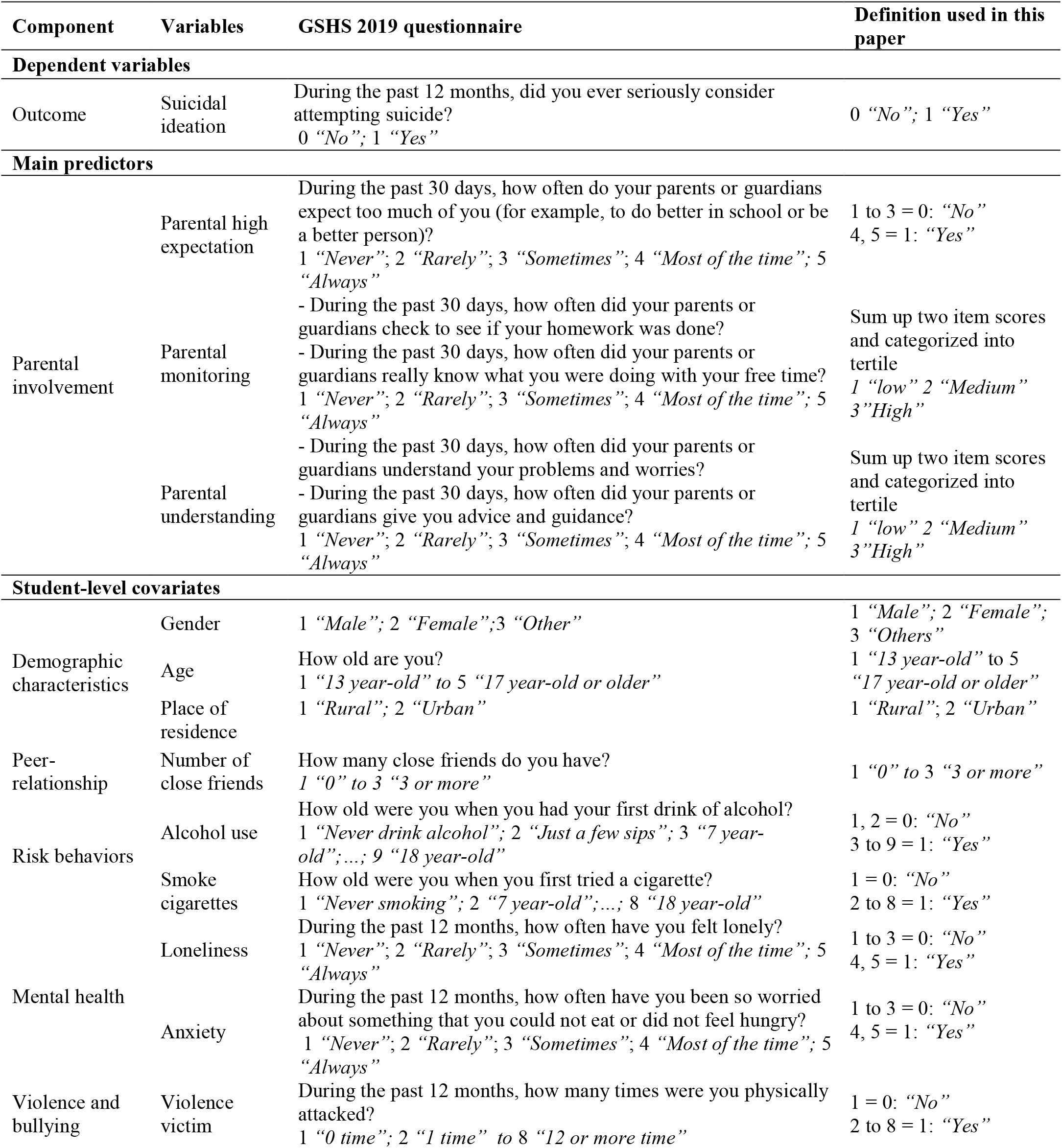
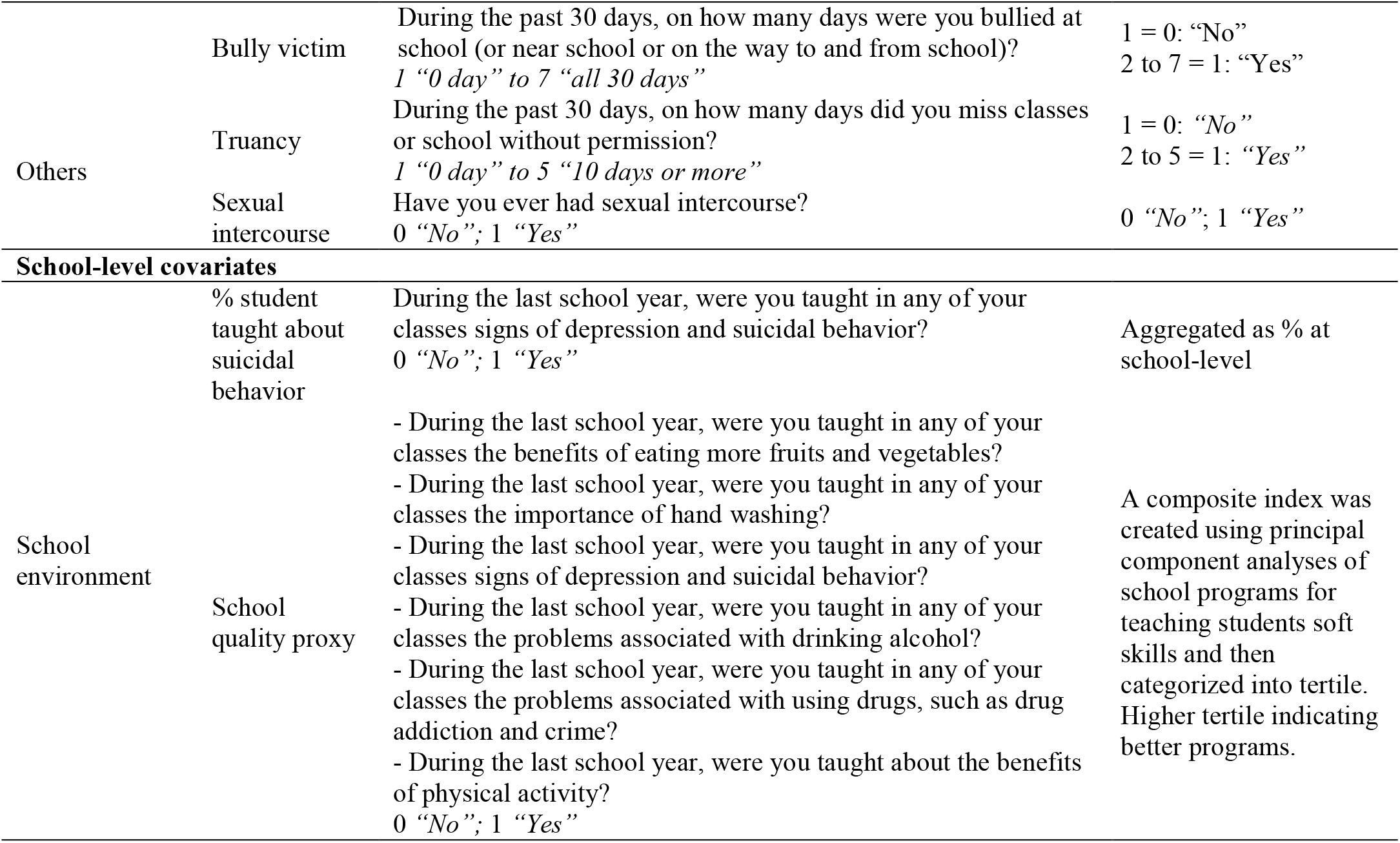
Variable definitions

#### Outcome variable

The outcome of interest was suicidal ideation (yes vs. no) within the last 12 months, which was assessed by asking students “During the past 12 months, did you ever seriously consider attempting suicide?”

#### Parental involvement

Parental involvement included three components: 1) high expectation, 2) parental monitoring, and 3) parental understanding. High expectation measured whether parents or guardians expected too much of students in a general aspect. Parental monitoring was assessed through whether parents or guardians checked homework and knew students’ activities in free time. Parental understanding was measured by the level of understanding of students’ problems and worries; and giving advice and guidance for students in the previous 30 days.

#### Other covariates

##### Student-level covariates

We assessed student-level variables included demographics characteristics (gender, age, place of residence); peer-relationship (number of close friends); risk behaviors (use of alcohol and cigarettes); mental health (loneliness and anxiety); sexual intercourse; truancy; violence and bullying.

##### School-level covariates

At the school level, we gathered two variables: 1) the percentage of students taught about suicidal behaviors and 2) school quality proxy measurement. We constructed the school quality proxy using principal component analysis (PCA) of school programs for teaching soft skills and then categorized them into tertile, with higher tertile indicates better programs (Vyas and Kumaranayake 2006).

### Data analysis

To summarize the data, we used frequency and weighted percentages for categorical variables; mean and 95% confidence interval (CI) for continuous variables. To compare the differences in the prevalence of suicidal ideation across groups, we calculated the weighted percentage and 95% CI of suicidal ideation stratified by students’ and schools’ characteristics. The weighted estimates accounted for sample weights, clustering of school, and stratification.

To evaluate the relationship between parental involvement and youth suicidal ideation, we fitted a series of 2-level random intercept logistic regression models with students at level-1, and school at level-2. Multilevel modeling is a statistical technique that provides an efficient framework for the complex survey design and accounts for the variation in hierarchical data (Goldstein 2010; Steele 2008).

The first model was a null model with no independent variable to serve as a base model (Model 1).

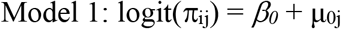

Where, β_0_ presents the median log odds of suicidal ideation probability of students across all schools. The term μ_0j_ is the school-specific residual that presents the deviate of each school from the national median log odds suicidal ideation. These school-specific residuals are assumed to be normally distributed (i.e., μ_0j_ ∼ N(0, σ^2^_μ0_)). For logistic models, the variance at the individual-level is assumed to be a function of the binomial distribution (i.e., pi^2^/3 = 3.29) (Goldstein 2010); thus, the variance partition coefficient (VPC) can be calculated as VPC = σ^2^_μ0_/(σ^2^_μ0_ + 3.29). The VPC indicates the proportion of total variance of suicidal ideation that is explained by the variation between schools.

In Model 2, we included three main predictors indicated parental involvement (denoted as PI) (i.e., parental high expectation, parental monitoring, and parental understanding). In model 3, we controlled for all other student-level covariates (denoted as X).

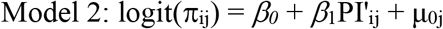

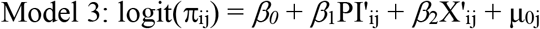

In Model 4, we added school-level variables (denoted as SL), including the percentage of students taught about symptoms of mental health and suicidal behaviors, and the school quality proxy variable.

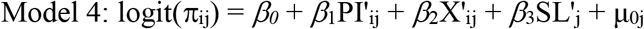

For each model, we calculated the proportion of variance of suicidal ideation explained by the added factors (i.e., % explained) by subtracting the variance of the simpler model to the model with more terms and then converted to percentage. We use odds ratio (OR) and 95% CI to report the modeling results, the likelihood ratio tests to compare the likelihood between logistic models with and without random effects. A significance level of 0.05 was used for all statistical tests. All analyses were carried out using Stata v16 SE (StataCorp, College Station, TX, USA).

## Ethical approval

All procedures performed in this study were in accordance with the ethical standards of the Institution Review Board of Hanoi University of Public Health (IRB decision No. 421/2019/YTCC-HD3, dated: 06/08/2019). Written informed consents were obtained from all participant’s parents/guardians before the study. All survey participants were voluntary, anonymous, and confidential. The students had the right to withdraw from the study or refuse to answer any specific questions in the questionnaire without any consequences.

## Results

### Participants’ characteristics

Table 2 describes the study participants’ characteristics. More than half of the participants were females (53.4%) and lived in rural areas (63.5%). Nearly two-thirds of the participants had three or more close friends (62.5%). The percentages of students drinking alcohol and smoking cigarettes in the past 12 months were 44.3% and 8.3%, respectively. A small number of participants experienced loneliness (12.6%) and anxiety (6.3%). About 15% did truant, while 5.2% experienced sexual intercourse. About 10.4% of students were violence victims and 5.6% was bully victims during the past 12 months. Regarding the school environment, 21.1% of students were taught about suicidal behaviors in school and 41.5% participated in schools with high quality scale. Parental involvement was moderate, with 54.7% of participants experiencing high parental expectation, 33.8% having high parental monitoring, and 27.7% receiving high parental understanding.

**Table 2.** Participants’ characteristics among in-school adolescents in Vietnam

|  | Participant characteristics |  |
| --- | --- | --- |
|  | Unweighted n | Weighted % |
|  | 7796 |  |
| **Suicidal ideation** |  |  |
| No | 6,400 | 84.4 |
| Yes | 1,386 | 15.6 |
| **Parental involvement** |  |  |
| **Parental high expectation** |  |  |
| No | 3,497 | 45.3 |
| Yes | 3,960 | 54.7 |
| **Parental monitoring** |  |  |
| Low | 2,899 | 34.7 |
| Medium | 2,527 | 31.5 |
| High | 2,341 | 33.8 |
| **Parental understanding** |  |  |
| Low | 3,373 | 42.0 |
| Medium | 2,438 | 30.2 |
| High | 1,983 | 27.7 |
| **Student-level variables** |  |  |
| **Gender** |  |  |
| Male | 3,572 | 45.4 |
| Female | 4,118 | 53.4 |
| Others | 106 | 1.2 |
| **Age** |  |  |
| 13 | 1,037 | 16.8 |
| 14 | 1,544 | 30.3 |
| 15 | 1,696 | 19.9 |
| 16 | 1,676 | 16.0 |
| 17+ | 1,843 | 17.1 |
| **Place of residence** |  |  |
| Rural | 3,878 | 63.5 |
| Urban | 3,918 | 36.5 |
| **Number of close friends** |  |  |
| 0 | 795 | 8.9 |
| 1 | 1,094 | 13.1 |
| 2 | 1,188 | 15.5 |
| ≥3 | 4,621 | 62.5 |
| **Drinking alcohol** |  |  |
| No | 4,020 | 55.7 |
| Yes | 3,731 | 44.3 |
| **Smoking cigarettes** |  |  |
| No | 7,042 | 91.7 |
| Yes | 731 | 8.3 |
| **Loneliness** |  |  |
| No | 6,660 | 87.4 |
| Yes | 1,133 | 12.6 |
| **Anxiety** |  |  |
| No | 7,222 | 93.7 |
| Yes | 564 | 6.3 |
| **Truancy** |  |  |
| No | 6,389 | 84.9 |
| Yes | 1,211 | 15.1 |
| **Sexual intercourse** |  |  |
| No | 7,275 | 94.8 |
| Yes | 477 | 5.2 |
| **Violence victim** |  |  |
| No | 6,971 | 89.6 |
| Yes | 822 | 10.4 |
| **Bully victim** |  |  |
| No | 7,346 | 94.4 |
| Yes | 445 | 5.6 |
| **School-level variables** |  |  |
| **Percentage of students taught about suicidal behavior, mean (95% CI)** |  | 21.1 (18.6–23.7) |
| **School quality proxy** |  |  |
| 1<sup>st</sup> tertile | 2,623 | 22.8 |
| 2<sup>nd</sup> tertile | 2,599 | 35.7 |
| 3<sup>rd</sup> tertile | 2,574 | 41.5 |
| CI: confidence interval |  |  |

### Relationship between parental involvement and suicidal ideation

Figure 2 illustrates the prevalence of suicidal ideation by parental involvement. Overall, 15.6% (95% CI: 14.2–17.1) of in-school adolescents had suicidal ideation at least once in the previous 12 months. Compared to students with low parental expectation, those with high parental expectation had higher prevalence of suicidal ideation (13.4% vs. 17.8%). In contrast, participants with medium and high levels of parental monitoring and understanding were less likely to have suicidal ideation compared to those with a low level (9–15% vs. 22%).

**Fig. 2.**
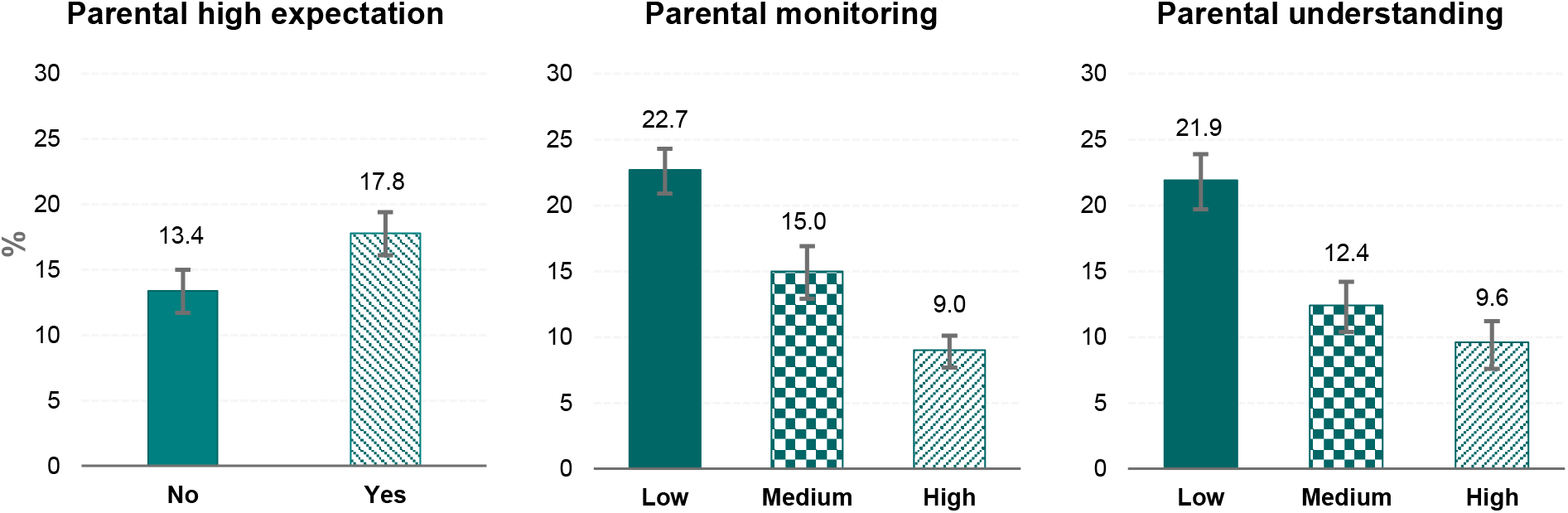
Prevalence of suicidal ideation by parental involvement among in-school adolescents in Vietnam.

### Factors related to suicidal ideation

Table 3 shows the multivariable models of factors related to suicidal ideation among in-school adolescents in Vietnam. In all models, the likelihood ratio tests compared between logistic models with and without random effects showed significant results, which indicated the existence of the clustering effects among students across schools.

**Table 3.** Multivariable models of factors related to suicide ideation among in-school adolescents in Vietnam

|  | Model 1 |  | Model 2 |  | Model 3 |  | Model 4 |  |
| --- | --- | --- | --- | --- | --- | --- | --- | --- |
|  | OR | 95% CI | OR | 95% CI | OR | 95% CI | OR | 95% CI |
| **Fixed part** |  |  |  |  |  |  |  |  |
| **Intercept** | 0.20*** | (0.18–0.23) | 0.29*** | (0.25–0.34) | 0.15*** | (0.10–0.21) | 0.16*** | (0.10–0.24) |
| **Parental high expectation**<br>(Ref: No) |  |  |  |  |  |  |  |  |
| Yes |  |  | 1.58*** | (1.39–1.79) | 1.39*** | (1.21–1.60) | **1.39\*\*\*** | **(1.21–1.61)** |
| **Parental monitoring**<br>(Ref: Low) |  |  |  |  |  |  |  |  |
| Medium |  |  | 0.74*** | (0.64–0.85) | 0.86 | (0.73–1.01) | 0.86 | (0.73–1.01) |
| High |  |  | 0.49*** | (0.40–0.58) | 0.63*** | (0.52–0.77) | **0.63\*\*\*** | **(0.52–0.77)** |
| **Parental understanding**<br>(Ref: Low) |  |  |  |  |  |  |  |  |
| Medium |  |  | 0.56*** | (0.48–0.65) | 0.64*** | (0.54–0.75) | **0.64\*\*\*** | **(0.54–0.75)** |
| High |  |  | 0.52*** | (0.43–0.63) | 0.60*** | (0.49–0.74) | **0.60\*\*\*** | **(0.49–0.73)** |
| **Gender (Ref: Male)** |  |  |  |  |  |  |  |  |
| Female |  |  |  |  | 1.68*** | (1.45–1.94) | **1.69\*\*\*** | **(1.46–1.95)** |
| Other |  |  |  |  | 2.29*** | (1.42–3.70) | **2.34\*\*\*** | **(1.45–3.77)** |
| **Age (Ref: 13)** |  |  |  |  |  |  |  |  |
| 14 |  |  |  |  | 0.85 | (0.67–1.08) | 0.85 | (0.67–1.09) |
| 15 |  |  |  |  | 0.85 | (0.65–1.10) | 0.80 | (0.62–1.04) |
| 16 |  |  |  |  | 0.79 | (0.60–1.03) | 0.73* | (0.56–0.95) |
| 17+ |  |  |  |  | 0.86 | (0.66–1.12) | 0.80 | (0.61–1.04) |
| **Place of residence**<br>(Ref: Rural) |  |  |  |  |  |  |  |  |
| Urban |  |  |  |  | 1.44*** | (1.19–1.73) | **1.29\*\*** | **(1.07–1.57)** |
| **Number of close friends**<br>(Ref: 0) |  |  |  |  |  |  |  |  |
| 1 |  |  |  |  | 0.72** | (0.56–0.91) | **0.72\*\*** | **(0.56–0.91)** |
| 2 |  |  |  |  | 0.90 | (0.71–1.14) | 0.90 | (0.71–1.14) |
| ≥3 |  |  |  |  | 0.56*** | (0.46–0.69) | **0.56\*\*\*** | **(0.46–0.69)** |
| **Alcohol use (Ref: No)** |  |  |  |  |  |  |  |  |
| Yes |  |  |  |  | 1.48*** | (1.28–1.71) | **1.48\*\*\*** | **(1.28–1.71)** |
| **Smoke cigarettes**<br>(Ref: No) |  |  |  |  |  |  |  |  |
| Yes |  |  |  |  | 1.29* | (1.03–1.60) | **1.28\*** | **(1.02–1.59)** |
| **Loneliness (Ref: No)** |  |  |  |  |  |  |  |  |
| Yes |  |  |  |  | 2.25*** | (1.90–2.67) | **2.25\*\*\*** | **(1.90–2.67)** |
| **Anxiety (Ref: No)** |  |  |  |  |  |  |  |  |
| Yes |  |  |  |  | 2.98*** | (2.40–3.69) | **3.01\*\*\*** | **(2.43–3.74)** |
| **Truancy (Ref: No)** |  |  |  |  |  |  |  |  |
| Yes |  |  |  |  | 1.17 | (0.97–1.39) | 1.18 | (0.98–1.41) |
| **Sexual intercourse (Ref: No)** |  |  |  |  |  |  |  |  |
| Yes |  |  |  |  | 1.03 | (0.78–1.35) | 1.02 | (0.78–1.34) |
| **Violence victim (Ref: No)** |  |  |  |  |  |  |  |  |
| Yes |  |  |  |  | 1.60*** | (1.30–1.96) | **1.59\*\*\*** | **(1.29–1.96)** |
| **Bully victim (Ref: No)** |  |  |  |  |  |  |  |  |
| Yes |  |  |  |  | 1.50** | (1.16–1.93) | **1.48\*\*** | **(1.15–1.92)** |
| **School-level variables** |  |  |  |  |  |  |  |  |
| **Percentage of students taught about suicidal behavior** |  |  |  |  |  |  | 1.01 | (1.00–1.02) |
|  | OR | 95% CI | OR | 95% CI | OR | 95% CI | OR | 95% CI |
| **School quality proxy** |  |  |  |  |  |  |  |  |
| *(Ref: 1<sup>st</sup> tertile)* |  |  |  |  |  |  |  |  |
| 2<sup>nd</sup> tertile |  |  |  |  |  |  | 0.88 | (0.70–1.11) |
| 3<sup>rd</sup> tertile |  |  |  |  |  |  | **0.68\*\*** | **(0.53–0.86)** |
| **Random part** |  |  |  |  |  |  |  |  |
| Variance estimate | 0.14 | (0.08–0.23) | 0.12 | (0.07–0.21) | 0.08 | (0.04–0.17) | 0.07 | (0.03–0.15) |
| VPC (%)<sup>§</sup> | 4.08 |  | 3.5 |  | 2.5 |  | 2.0 |  |
| % Explained<sup>†</sup> | -- |  | 14.1 |  | 29.8 |  | 22.2 |  |
| LR test<sup>‡</sup> | $\chi^2 = 61.9, p < 0.001$ | | $\chi^2 = 44.5, p < 0.001$ | | $\chi^2 = 19.5, p < 0.001$ | | $\chi^2 = 13.4, p < 0.001$ | |

CI: Confidence interval; LR: Likelihood ratio; OR: Odds ratio; VPC: Variance partition coefficient
\* $p < 0.05$ , \*\* $p < 0.01$ , \*\*\* $p < 0.001$
Model 1: A null 2-level random effects model, with students at level-1, and school at level-2
Model 2: Model 1 + main predictors (i.e., parental high expectation, parental monitoring, and parental understanding)
Model 3: Model 2 + all student-level covariates
Model 4: Model 3 + all school-level variables
<sup>§</sup>VPC calculated as: $[\sigma^2_{\mu 0}/(\sigma^2_{\mu 0} + 3.29)] \times 100$
<sup>†</sup>% Explained calculated as: $[(\sigma^2_{\text{Model N}} - \sigma^2_{\text{Model N} + 1})/\sigma^2_{\text{Model N}}] \times 100$
<sup>‡</sup>Likelihood ratio test of logistic model with random effects vs. logistic model with only fixed effect

In the base model without independent variable (Model 1), most variation of suicidal ideation attributed to the student level; the school-level only accounted for 4.08% of the variation. In Model 2, parental involvement variables were significantly associated with suicidal ideation. The inclusion of these variables explained 14.1% of the between-school variation of suicidal ideation. In Model 3, after controlling for student-level covariates, the odds of suicidal ideation were lower among students with high level of parental monitoring (OR: 0.63; 95% CI: 0.52–0.77) and with medium and high parental understanding (OR: 0.64; 95% CI: 0.54–0.75 and OR: 0.60; 95% CI: 0.49–0.73, respectively). In contrast, the odds of suicidal ideation were higher among adolescents with high parental expectation (OR: 1.42; 95% CI: 1.24–1.63). Compared to Model 2, additional student-level covariates explained 29.8% of variation at the school level.

In Model 4, the association between parental involvement and adolescent suicidal ideation remained the same after adjusting for school-level covariates. Schools at the 3^rd^ tertile of quality scale were associated with 32% lower odds of student suicidal ideation compared to school at 1^st^ tertile (OR: 0.68; 95% CI: 0.53–0.86). The percentage of students taught about suicidal behaviors at school did not relate to student suicidal ideation. Relative to Model 3, school-level indicators explained 22.2% between-school variation in suicidal ideation.

We also found associations between several individual-level covariates and suicidal ideation (Table 3). For example, we observed higher odds of suicidal ideation in females and other-gender students, those who lived in urban areas, and those who had risky behaviors (such as drinking alcohol or smoking cigarettes). Students who had any mental health symptoms or suffered from violence and bullying also had higher odds of suicidal ideation compared to their counterparts. In contrast, having more than two close friends was a significant protective factor compared to having no close friend.

Patterns of factors related to suicidal ideation were similar for male and female students. (Supplemental Table 3).

## Discussion

Suicidal ideation among adolescents is a public health concern; however, its link with parental involvement has not been well studied. We found that the prevalence of suicidal ideation among Vietnamese adolescents was 15.6%. Perceiving parental understanding and parental monitoring were sources of protection for adolescent suicidal ideation, while high parental expectation was associated with higher prevalence of suicidal ideation. Other factors associated with a higher risk of adolescent suicidal ideation included being a female or other-gender student, living in urban areas, involving in risky behaviors (alcohol use or smoking cigarettes), having mental health issues (loneliness or anxiety), having fewer close friends, and experiencing bullying at school or violence. Our findings serve as important evidence for developing effective prevention and action strategies to mitigate the burden of youth suicide in Vietnam.

The prevalence of suicidal ideation among Vietnamese youth in 2019 slightly reduces compared to data in 2013 (16.9%) (Nguyen et al. 2019). Adolescent suicidal ideation in Vietnam occurs at a similar rate compared to 49 other LMICs (average prevalence of 15.3%) (Page and West 2011). This rate is lower than findings from few other African countries; i.e. 31% in Zambia (Muula et al. 2007) and 22% in Uganda (Rudatsikira et al. 2007), but higher than other South East Asian countries (Thailand 8.8%, Myanmar 0.7%, Indonesia 4.0%) (Peltzer and Pengpid 2012, 2017). Indeed, Vietnam ranked second in the prevalence of suicidal ideation among ASEAN countries (Peltzer and Pengpid 2017). This high rate of suicidal ideation highlights the need for specific policies and effective solutions from health professionals as well as government and non-governmental organizations to mitigate the burden in adolescent suicide.

The influence of parental involvement on youth suicide rate is still controversial. Some researchers sustain that family unit is the single most important factor in understanding suicide (Diekstra et al. 1989). A report by the World Health Organization also identified family context (quality parenting style) as one of the main protective factors of youth suicide. However, while “good” parenting can be salubrious, “poor” parenting may exacerbate children’s mental and psychological problems, hence increases the odds of youth suicidal ideation. A previous finding shows that “overly controlling” parenting style adds to youth suicide outcomes (Goschin et al. 2013). In our study, we found that parental understanding and monitoring may decrease suicidal thoughts among adolescents in Vietnam. We also found a reverse effect of high parental expectation on suicidal ideation, which is consistent with previous evidence (Lee et al. 2006). High parental expectation for children’s academic performance may lead to excessive pressure, and high level of parental control (Roth et al. 2009), which can increase the risk of mental health problems and suicidal ideation.

Modern trends emphasize adolescents’ competence and independence, but parental support still plays a vital role in improving children’s social functions and promoting their mental health (Moretti and Peled 2004). Results from a study in Hong Kong, which is partially similar to Vietnam in culture, indicated that a high degree of understanding and monitoring from parents reduced the likelihood of mental issues (Klemera et al. 2017; Law and Shek 2013). Compared to the Western cultures, the Asian culture (including Vietnam) generally emphasizes obedience, respect toward elders, and parental authority (i.e., parents tend to use more commands and attempt to control their children) (Russell ST et al. 2010). Negative feelings regarding lower level of independence might link to suicidal ideation among adolescents (Gouveia-Pereira et al. 2014). However, evidence regarding causality from parental involvement to children suicidal ideation remains insufficient. This study revealed both positive and negative impacts of parental involvement on suicidal ideation. We suggest including parental involvement and family-related factors when developing interventions to prevent suicidal ideation in adolescents.

Our findings indicate that both school-level and student-level factors are associated with students’ suicidal ideation. We found 21.1% of students were taught about suicidal behaviors, however, this was not significantly associated to suicidal ideation. Currently, teaching about suicidal behaviors in Vietnam is not officially in the school curriculum, but only introduced in some extra activities, therefore, the time and quality of training might be not sufficient. Consistent with findings from other studies, we found that high school quality was negatively correlated with suicidal ideation. The between-school variations from multilevel models were relatively low (the VPC was 4.08%), which aligned with findings from previous studies (Sellström and Bremberg 2006). However, the school-effect still plays a vital role in developing policies to prevent youth suicidal ideation. Together, these findings highlight the need for the appropriate training programs which focus more on quality of the content to provide essential knowledge of suicidal behaviors for adolescents. For student-level factors, we confirmed the association between increased suicidal ideation and poor mental health and being victims of bullying or violence as shown in current literature (Itani et al. 2017; Page and West 2011; Peltzer and Pengpid 2015, 2017).

Adolescents have the power to transform the future of the country; improving physical and mental health of the young is an integral part of reaching the Sustainable Development Goals (SDG) (i.e., SDG 3: Ensure healthy lives and promote well-being for all at all ages) (World Health Organization). Parents and teachers play important roles in assessing youth suicidal risk since adolescents usually spend a substantial amount of time at school and at home. Therefore, training school staff and parents for warning signs of adolescents’ suicidal behaviors is essential. Our findings from both levels contribute to important practical implications for suicide prevention and intervention. We recommend integration of supportive mental health services and engagement of school community and parents through student behaviors.

### Strengths and limitations

This is one of very few studies to investigate the current status of youth suicidal behaviors in Vietnam, using nationally representative data with large sample size. We used a strong conceptual framework to provide an integrated theoretical and analytical underpinning that articulates the research foci and links between parental involvement and children’s suicide ideation. By using multilevel models, we are able account for the hierarchical data structure that students are nested within schools. This technique also allowed us to simultaneously evaluate the variation in suicidal ideation at different levels (i.e., student level and school level) and the effects of each level’s factors, which increases the explainability of our findings.

We acknowledge some limitations when interpreting findings of this study. First, the GSHS 2019 used the single-item, self-report measure to assess suicidal ideation. The prevalence of suicidal ideation, therefore, might be underestimated. Second, although the GSHS are large, nationally representative, and contained rich data on adolescent and youth behaviors, several important factors known to be associated with suicidal ideation were not available. For example, the survey did not assess household social-economic status, parental education, and mental health history of family members. Third, the cross-sectional nature of data prevented us from inferring the causal direction between youth suicide and parental involvement conclusively.

## Conclusions

Suicidal ideation is prevalent among school-going adolescents in Vietnam as an example of a LMIC. Parental involvement shows both positive and negative impacts on adolescent suicidal ideation - high parental expectation increases suicidal risk while parental understanding and parental monitoring reduce the risk. This study can inform policymakers and other potential stakeholders and guide interventions that target parent-child bonding and other factors at student- and school-level in the efforts to mitigate the burden of suicidal behaviors and achieve SDG on well-being for adolescents.

## Data Availability

The datasets analyzed for the current study are not publicly available but are available upon reasonable request

## Declarations

### Compliance with Ethical Standards

#### Funding

The 2019 Global School-based Student Health Survey was conducted with financial support from the World Health Organization. The authors received no funding for the data analysis, data interpretation, manuscript writing, authorship, and/or publication of this article.

#### Consent to participate

Written informed consents were obtained from all participants’ parents/guardians before the study.

## Authors’ contributions

KQL conceived of the study, performed the statistical analyses, interpreted the results, and drafted the manuscript; NHP interpreted the results, reviewed and revised the manuscript; HTNA collected data and drafted the manuscript; TTTH coordinated the data collection, collected data, reviewed and revised the manuscript; NHNV, PQT, NHP coordinated the data collection and collected data; LPA collected data, reviewed and revised the manuscript; KP, MT, NTL, PTQN, LVT, TQB, LDMA, HVM reviewed and revised the manuscript. All authors read and approved the final manuscript.

## Acknowledgments

The authors would like to thank all students who participated in this study as well as the individuals and institutions that made this research possible: Departments of Education and Training from selected provinces, principals and teachers from 81 schools who helped us to prepare for the data collection. Associate Prof. Tran Dac Phu and Dr. Truong Dinh Bac from the General Department of Preventive Medicine – Ministry of Health; Mr. Nguyen Thanh De and Mr. Le Manh Hung from the Ministry of Education and Training; Ms. Leanne Riley from World Health Organization; Ms. Veronica Lea, Ms. Curtis Blanton and Mr. Timothy McManus from the US Centers for Disease Control and Prevention, and Mr. Cao Huu Quang from Hanoi University of Public Health.

