## Supplemental materials for "Relationships Between Parental Involvement and Suicidal Ideation among In-school Adolescents in Vietnam: A Multilevel Analysis of the Global School-Based Student Health Survey 2019"

### Supplemental Table 1: Participants' characteristics, stratified by gender

| | Male | | Female | | Others | |
|---|---|---|---|---|---|---|
| **Characteristics** | **n** | **weighted %** | **n** | **weighted %** | **n** | **weighted %** |
| **N** | 3,572 | | 4,118 | | 106 | |
| **High expectation** | | | | | | |
| No | 1,529 | 43.1 | 1,910 | 47.1 | 58 | 48.3 |
| Yes | 1,879 | 56.9 | 2,035 | 52.9 | 46 | 51.7 |
| **Parental monitoring** | | | | | | |
| Low | 1,222 | 32.4 | 1,626 | 36.4 | 51 | 43.6 |
| Medium | 1,151 | 30.8 | 1,343 | 32.1 | 33 | 30.8 |
| High | 1,182 | 36.8 | 1,138 | 31.5 | 21 | 25.6 |
| **Parental understanding** | | | | | | |
| Low | 1,463 | 40.1 | 1,852 | 43.5 | 58 | 53.3 |
| Medium | 1,145 | 30.3 | 1,262 | 30.3 | 31 | 25.0 |
| High | 963 | 29.6 | 1,003 | 26.3 | 17 | 21.7 |
| **Age** | | | | | | |
| 13 | 441 | 15.7 | 583 | 17.6 | 13 | 23.8 |
| 14 | 684 | 28.6 | 839 | 31.9 | 21 | 19.3 |
| 15 | 818 | 21.7 | 854 | 18.3 | 24 | 23.8 |
| 16 | 753 | 16.1 | 893 | 15.7 | 30 | 20.5 |
| 17+ | 876 | 17.9 | 949 | 16.5 | 18 | 12.6 |
| **Place of residence** | | | | | | |
| Rural | 1,755 | 62.9 | 2,078 | 64.1 | 45 | 61.6 |
| Urban | 1,817 | 37.1 | 2,040 | 35.9 | 61 | 38.4 |
| **Number of close friends** | | | | | | |
| 0 | 353 | 8.5 | 425 | 9.0 | 17 | 21.6 |
| 1 | 397 | 10.4 | 678 | 15.2 | 19 | 19.7 |
| 2 | 457 | 13.0 | 715 | 17.8 | 16 | 10.8 |
| ≥3 | 2,318 | 68.2 | 2,251 | 58.0 | 52 | 47.8 |
| **Drinking alcohol** | | | | | | |
| No | 1,730 | 53.6 | 2,257 | 58.0 | 33 | 29.4 |
| Yes | 1,818 | 46.4 | 1,841 | 42.0 | 72 | 70.6 |
| **Smoking cigarettes** | | | | | | |
| No | 3,032 | 86.7 | 3,922 | 96.0 | 88 | 86.0 |
| Yes | 525 | 13.3 | 188 | 4.0 | 18 | 14.0 |
| **Loneliness** | | | | | | |
| No | 3,088 | 88.3 | 3,501 | 87.2 | 71 | 61.5 |
| Yes | 481 | 11.7 | 617 | 12.8 | 35 | 38.5 |
| **Anxiety** | | | | | | |
| No | 3,367 | 95.2 | 3,766 | 92.7 | 89 | 84.2 |
| Yes | 198 | 4.8 | 350 | 7.3 | 16 | 15.8 |
| **Truancy** | | | | | | |
| No | 2,834 | 82.6 | 3,473 | 86.9 | 82 | 85.6 |
| Yes | 634 | 17.4 | 557 | 13.1 | 20 | 14.4 |
| **Sexual intercourse** | | | | | | |
| No | 3,272 | 93.1 | 3,916 | 96.4 | 87 | 83.6 |
| Yes | 281 | 6.9 | 178 | 3.6 | 18 | 16.4 |
| **Violence victim** | | | | | | |
| No | 3,054 | 84.6 | 3,831 | 93.9 | 86 | 83.2 |
| Yes | 517 | 15.4 | 285 | 6.1 | 20 | 16.8 |
| **Bully victim** | | | | | | |
| No | 3,339 | 93.4 | 3,919 | 95.5 | 88 | 85.5 |
| Yes | 228 | 6.6 | 199 | 4.5 | 18 | 14.5 |

**Supplemental Table 2. Suicide ideation prevalence among in-school adolescents in Vietnam, stratified by gender**

| | Male | | Female | | Others | |
|---|---|---|---|---|---|---|
| | % | 95% CI | % | 95% CI | % | 95% CI |
| **Overall** | **11.9** | **10.6, 13.3** | **18.4** | **16.9, 20.0** | **35.6** | **22.1, 52.0** |
| **High expectation** | | | | | | |
| No | 10.7 | 9.0, 12.8 | 15.1 | 13.1, 17.4 | 28.0 | 15.5, 45.3 |
| Yes | 13.3 | 11.8, 14.9 | 21.4 | 19.6, 23.4 | 41.7 | 24.6, 61.1 |
| **Parental monitoring** | | | | | | |
| Low | 17.4 | 14.9-20.3 | 26.2 | 23.5-29.1 | 44.6 | 28.5-62.0 |
| Medium | 11.9 | 9.9-14.4 | 17.1 | 14.3-20.2 | 33.4 | 18.0-53.4 |
| High | 7.1 | 5.4-9.3 | 10.7 | 8.9-12.7 | 23.4 | 7.2-54.8 |
| **Parental understanding** | | | | | | |
| Low | 16.2 | 13.8-19.1 | 25.9 | 23.3-28.7 | 40.6 | 24.1-59.5 |
| Medium | 9.2 | 7.2-11.6 | 14.7 | 12.7-16.9 | 33.9 | 14.9-59.9 |
| High | 8.8 | 6.8-11.3 | 10.1 | 7.7-13.1 | 25.4 | 9.2-53.5 |
| **Age** | | | | | | |
| 13 | 10.8 | 8.2, 14.1 | 18.1 | 14.4, 22.5 | 45.4 | 14.6, 80.2 |
| 14 | 9.5 | 7.3, 12.4 | 16.7 | 13.8, 20.2 | 33.9 | 14.6, 60.5 |
| 15 | 12.0 | 9.5, 15.1 | 15.8 | 12.1, 20.3 | 33.2 | 16.0, 56.3 |
| 16 | 13.2 | 10.6, 16.4 | 21.0 | 17.3, 25.3 | 32.5 | 16.8, 53.4 |
| 17+ | 15.2 | 12.6, 18.3 | 22.0 | 19.0, 25.4 | 29.5 | 11.6, 57.3 |
| **Place of residence** | | | | | | |
| Rural | 10.2 | 8.8, 11.9 | 15.8 | 13.8, 18.0 | 31.0 | 12.5, 58.4 |
| Urban | 14.7 | 12.7, 17.0 | 22.9 | 20.5, 25.5 | 43.1 | 30.8, 56.3 |
| **Number of close friends** | | | | | | |
| 0 | 23.8 | 19.7, 28.4 | 30.0 | 25.2, 35.4 | 42.2 | 22.1, 65.1 |
| 1 | 15.1 | 11.1, 20.2 | 18.7 | 14.8, 23.4 | 19.7 | 6.2, 48.0 |
| 2 | 14.1 | 10.8, 18.1 | 22.9 | 19.2, 27.1 | 40.3 | 19.4, 65.4 |
| ≥3 | 9.3 | 7.8, 11.2 | 14.8 | 13.2, 16.7 | 39.0 | 20.1, 61.7 |
| **Alcohol use** | | | | | | |
| No | 8.8 | 7.3, 10.6 | 12.8 | 11.0, 14.9 | 37.7 | 20.0, 59.4 |
| Yes | 15.5 | 13.4, 17.7 | 25.7 | 22.9, 28.7 | 35.0 | 19.8, 54.0 |
| **Smoke cigarettes** | | | | | | |
| No | 10.9 | 9.6, 12.4 | 17.4 | 15.9, 18.9 | 34.5 | 22.0, 49.7 |
| Yes | 18.3 | 14.7, 22.5 | 41.0 | 29.7, 53.4 | 42.2 | 15.3, 74.7 |
| **Loneliness** | | | | | | |
| No | 9.2 | 8.2, 10.3 | 15.0 | 13.5, 16.5 | 27.5 | 15.4, 44.2 |
| Yes | 32.3 | 26.1, 39.2 | 41.6 | 36.5, 46.9 | 48.6 | 24.8, 73.0 |
| **Anxiety** | | | | | | |
| No | 10.5 | 9.2, 11.9 | 15.7 | 14.1, 17.5 | 28.9 | 16.7, 45.0 |
| Yes | 39.4 | 29.7, 50.1 | 51.7 | 44.2, 59.1 | 72.6 | 35.8, 92.6 |
| **Truancy** | | | | | | |
| No | 11.1 | 9.6, 12.7 | 17.1 | 15.7, 18.6 | 36 | 21.2, 54.1 |
| Yes | 15.2 | 12.9, 17.9 | 25.4 | 20.1, 31.6 | 25 | 12.1, 44.6 |
| **Sexual intercourse** | | | | | | |
| No | 11.5 | 10.2, 12.9 | 18.1 | 16.6, 19.7 | 37.3 | 21.7, 56.0 |
| Yes | 17.2 | 13.0, 22.3 | 24.4 | 16.9, 34.0 | 27.9 | 7.4, 65.2 |
| **Violence victim** | | | | | | |
| No | 10.9 | 9.8, 12.1 | 17.0 | 15.6, 18.5 | 35.8 | 21.8, 52.9 |
| Yes | 17.4 | 13.5, 22.0 | 39.2 | 33.3, 45.5 | 34.6 | 14.3, 62.6 |
| **Bully victim** | | | | | | |
| No | 10.9 | 9.8, 12.2 | 17.4 | 15.9, 19.0 | 36.8 | 22.7, 53.6 |
| Yes | 24.8 | 18.2, 32.7 | 38.9 | 25.4, 54.4 | 28.5 | 11.8, 54.3 |

**Supplemental Table 3. Multivariable models of factors related to suicide ideation among in-school adolescents in Vietnam, stratified by gender**

| | Male | | Female | |
|---|---|---|---|---|
| | **OR** | **95% CI** | **OR** | **95% CI** |
| **Fixed part** | | | | |
| **Intercept** | 0.21*** | (0.12, 0.37) | 0.24*** | (0.14, 0.41) |
| **High expectation** *(Ref: No)* | | | | |
| Yes | 1.25* | (1.00, 1.55) | 1.48*** | (1.23, 1.77) |
| **Parental monitoring** *(Ref: Low)* | | | | |
| Medium | 0.85 | (0.65, 1.09) | 0.88 | (0.71, 1.08) |
| High | 0.60** | (0.44, 0.82) | 0.66** | (0.50, 0.86) |
| **Parental understanding** *(Ref: Low)* | | | | |
| Medium | 0.65** | (0.50, 0.85) | 0.63*** | (0.51, 0.77) |
| High | 0.77 | (0.57, 1.04) | 0.50*** | (0.38, 0.65) |
| **Age** *(Ref: 13)* | | | | |
| 14 | 0.72 | (0.48, 1.09) | 0.92 | (0.67, 1.25) |
| 15 | 0.87 | (0.59, 1.29) | 0.72 | (0.51, 1.02) |
| 16 | 0.66* | (0.44, 1.00) | 0.74 | (0.52, 1.05) |
| 17+ | 0.73 | (0.49, 1.08) | 0.79 | (0.56, 1.12) |
| **Place of residence** *(Ref: Rural)* | | | | |
| Urban | 1.28* | (1.01, 1.62) | 1.27 | (0.98, 1.66) |
| **Number of close friends** *(Ref: 0)* | | | | |
| 1 | 0.72 | (0.48, 1.07) | 0.73 | (0.53, 1.01) |
| 2 | 0.70 | (0.48, 1.04) | 0.99 | (0.72, 1.36) |
| ≥3 | 0.49*** | (0.36, 0.67) | 0.60*** | (0.46, 0.80) |
| **Alcohol use** *(Ref: No)* | | | | |
| Yes | 1.31* | (1.04, 1.65) | 1.64*** | (1.35, 1.97) |
| **Smoke cigarettes** *(Ref: No)* | | | | |
| Yes | 1.13 | (0.84, 1.52) | 1.75** | (1.22, 2.50) |
| **Loneliness** *(Ref: No)* | | | | |
| Yes | 2.65*** | (2.03, 3.45) | 1.98*** | (1.57, 2.49) |
| **Anxiety** *(Ref: No)* | | | | |
| Yes | 2.84*** | (1.98, 4.07) | 3.14*** | (2.38, 4.15) |
| **Truancy** *(Ref: No)* | | | | |
| Yes | 1.30 | (0.99, 1.70) | 1.13 | (0.88, 1.44) |
| **Sexual intercourse** *(Ref: No)* | | | | |
| Yes | 1.06 | (0.72, 1.55) | 0.94 | (0.62, 1.42) |
| **Violence victim** *(Ref: No)* | | | | |
| Yes | 1.30 | (0.97, 1.75) | 2.00*** | (1.47, 2.72) |
| **Bully victim** *(Ref: No)* | | | | |
| Yes | 1.46 | (0.99, 2.15) | 1.61** | (1.12, 2.31) |
| **School level variables** | | | | |
| **Percentage of students taught about suicidal behavior** | 1.01 | (1.00, 1.03) | 1.00 | (0.99, 1.02) |
| **School quality proxy** *(Ref: 1st tertile)* | | | | |
| 2nd tertile | 0.81 | (0.62, 1.07) | 0.98 | (0.72, 1.35) |
| 3rd tertile | 0.62** | (0.46, 0.84) | 0.73 | (0.52, 1.02) |
| **Random part** | | | | |
| Variance estimate | 0.0 | -- | 0.14 | 0.07, 0.29 |
| VPC (%)§ | -- | -- | 4.01 | |
| LR testΨ | $\chi$ = 0, p = 1.000 | | $\chi$ = 17.8, p < 0.001 | |

* *p < 0.05*, ** *p < 0.01*, *** *p < 0.001*

§VPC calculated as: $[\sigma^2_{\mu 0}/(\sigma^2_{\mu 0} + 3.29)]*100$

ΨLikelihood ratio test of logistic model with random effects vs. logistic model with only fixed effect

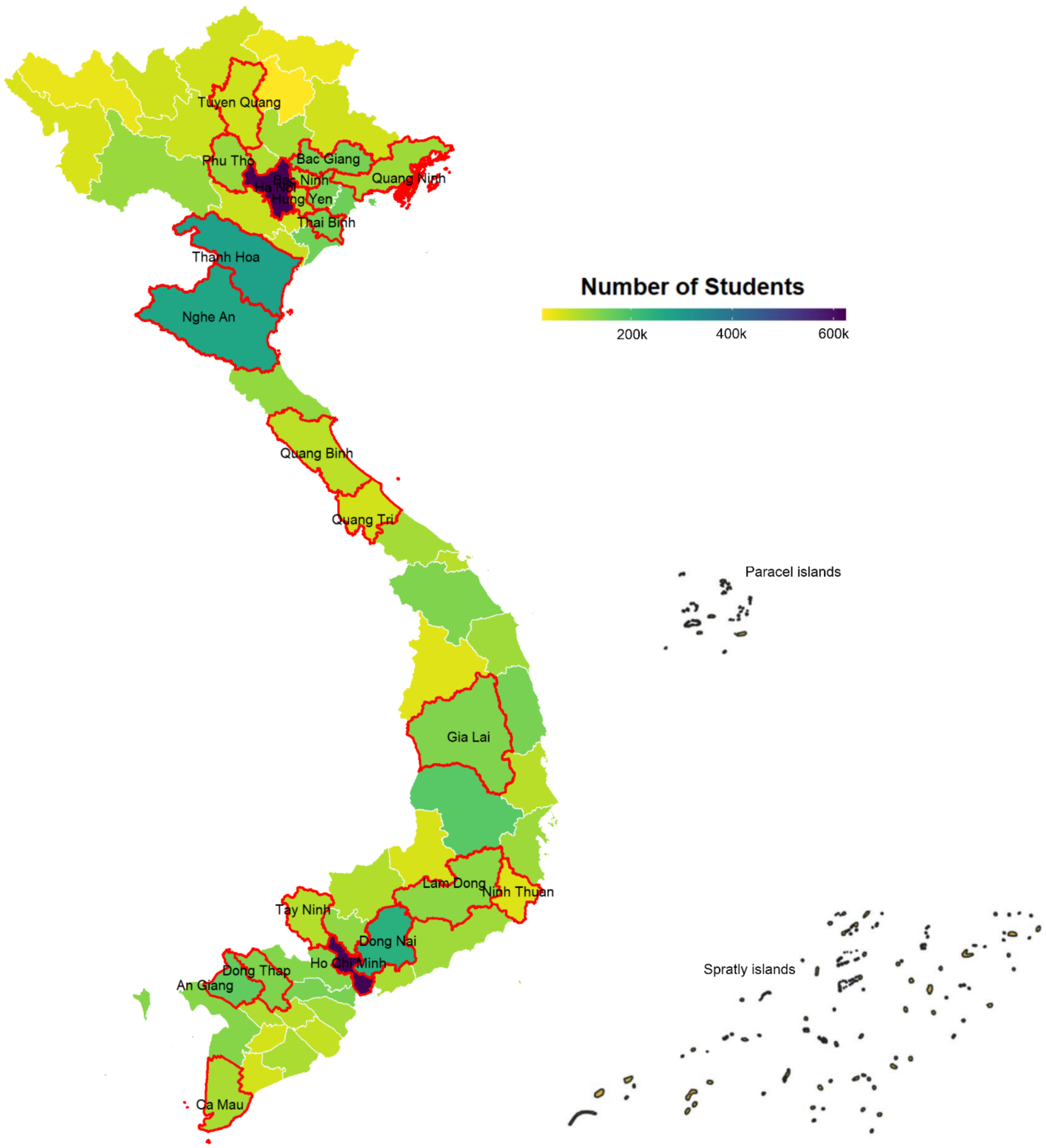


**Supplemental Figure 1: Selected provinces and cities for GSHS Vietnam 2019**
